# Evaluating Large Language Models on Medical Evidence Summarization

**DOI:** 10.1101/2023.04.22.23288967

**Authors:** Liyan Tang, Zhaoyi Sun, Betina Idnay, Jordan G Nestor, Ali Soroush, Pierre A. Elias, Ziyang Xu, Ying Ding, Greg Durrett, Justin Rousseau, Chunhua Weng, Yifan Peng

**Affiliations:** School of Information, The University of Texas at Austin, Austin, TX; Department of Population Health Sciences, Weill Cornell Medicine, New York, NY; Department of Biomedical Informatics, Columbia University, New York, NY; Department of Medicine, Columbia University, New York, NY; Department of Medicine, Massachusetts General Hospital, Boston, MA; Department of Computer Science, The University of Texas at Austin, Austin, TX; Departments of Population Health and Neurology, Dell Medical School, The University of Texas at Austin, Austin, TX

**Keywords:** Natural Language Processing, Medical Evidence Summarization, Large Language Model, Zero-shot, Human Evaluation, Error Types, Factual Consistency, Medical Harmfulness

## Abstract

Recent advances in large language models (LLMs) have demonstrated remarkable successes in zero- and few-shot performance on various downstream tasks, paving the way for applications in high-stakes domains. In this study, we systematically examine the capabilities and limitations of LLMs, specifically GPT-3.5 and ChatGPT, in performing zero-shot medical evidence summarization across six clinical domains. We conduct both automatic and human evaluations, covering several dimensions of summary quality. Our study has demonstrated that automatic metrics often do not strongly correlate with the quality of summaries. Furthermore, informed by our human evaluations, we define a terminology of error types for medical evidence summarization. Our findings reveal that LLMs could be susceptible to generating factually inconsistent summaries and making overly convincing or uncertain statements, leading to potential harm due to misinformation. Moreover, we find that models struggle to identify the salient information and are more error-prone when summarizing over longer textual contexts.

## 1 Introduction

Fine-tuned pre-trained models have been the leading approach in text summarization research, but they often require sizable training datasets which are not always available in specific domains, such as medical evidence in the literature. The recent success of zero- and few-shot prompting with large language models (LLMs) has led to a paradigm shift in NLP research [1–4]. The success of prompt-based models (GPT-3.5 [5], and recently ChatGPT [6]) brings new hope for medical evidence summarization, where the model can follow human instructions and summarize zero-shot without updating parameters. While recent work has analyzed and evaluated this strategy for news summarization [7] and biomedical literature abstract generation [8], there is no study yet on medical evidence summarization and evaluation. In this study, we conduct a systematic study of the potential and possible limitations of zero-shot prompt-based LLMs on medical evidence summarization using GPT-3.5 and ChatGPT models. We then explored their impact on the summarization of medical evidence findings in the context of evidence synthesis and meta-analysis.

## 2 Results

We made use of Cochrane Reviews obtained from the Cochrane Library and focused on six distinct clinical domains – Alzheimer’s disease, Kidney disease, Esophageal cancer, Neurological conditions, Skin disorders, and Heart failure. We collected around ten of the most recent reviews for each of these six domains. Domain experts verified each review to confirm that they have significant research objectives.

In our study, we tackle the single-document summarization setting where we focus on the abstracts of Cochrane Reviews. Different from other types of reviews, abstracts of Cochrane Reviews can be read as stand-alone documents [9]. They summarize the key methods, results and conclusions of the review. An abstract does not contain any information that is not in the main body of the review, and the overall messages should be consistent with the conclusions of the review. In addition, abstracts of Cochrane Reviews are freely available on the Internet (e.g., MEDLINE). As some readers may be unable to access the full review, abstracts may be the only source readers have to understand the review results.

Each abstract includes the Background, Research objectives, Search methods, Selection criteria, Data collection and analysis, Main results, and Author’s conclusions. The average length of the abstract we evaluated can be found in Table 1. We chose the *Author’s Conclusions* section, the last section of the abstract, as the human reference summary for the Cochrane Reviews in our study. This section contains the most salient details of the studies analyzed within the specific clinical context. Clinicians often consult the conclusions first when seeking answers to a clinical question, before deciding whether to read the full abstract and subsequently the entire study. It also allows the authors to interpret the evidence presented in the review, assess the strength of the evidence, and provide their own conclusions or recommendations concerning the efficacy and safety of the intervention under review.

**Table 1:** The number of words in the summarization dataset used for human evaluation.

| Domain | Count | Abstract | Main results | Conclusion |
| --- | --- | --- | --- | --- |
| Alzheimer’s disease | 10 | 678 | 449 | 114 |
| Kidney disease | 10 | 887 | 564 | 95 |
| Neurological conditions | 10 | 791 | 480 | 106 |
| Skin disorders | 9 | 1008 | 763 | 138 |
| Heart failure | 7 | 804 | 542 | 103 |
| Esophageal cancer | 7 | 632 | 397 | 119 |

We assessed the zero-shot performance of medical evidence summarization using two models: GPT-3.5 [5] (text-davinci-003) and ChatGPT [6]. To evaluate the models’ capabilities, we designed two distinct experimental setups. In the first setup, the models were given the entire abstract, excluding the *Author’s Conclusions* (ChatGPT-Abstract). In the second setup, the models received both the *Objectives* and the *Main Results* sections from the abstract as the input (ChatGPT-MainResult and GPT3.5-MainResult). Here, we chose Main Results as the input document because it includes the findings of all important benefit and harm outcomes. It also summarizes the impact of the risk of bias on trial design, conduct, and reporting. We did not evaluate GPT3.5-Abstract because our pilot study indicated that ChatGPT-MainResult performs generally better than ChatGPT-Abstract. In both settings, we used *[input*] + “*Based on the Objectives, summarize the above systematic review in four sentences*” as a prompt, emphasizing the importance of referring to the *Objectives* section for aspect-based summarization. We decided to summarize the review into four sentences since it is close to the length of human reference summaries on average.

Figure 1 presents a comparative analysis of summaries generated by ChatGPT-MainResult, ChatGPT-Abstract, and GPT3.5-MainResult across six clinical domains, as detailed in the Methods section.

**Fig. 1:**
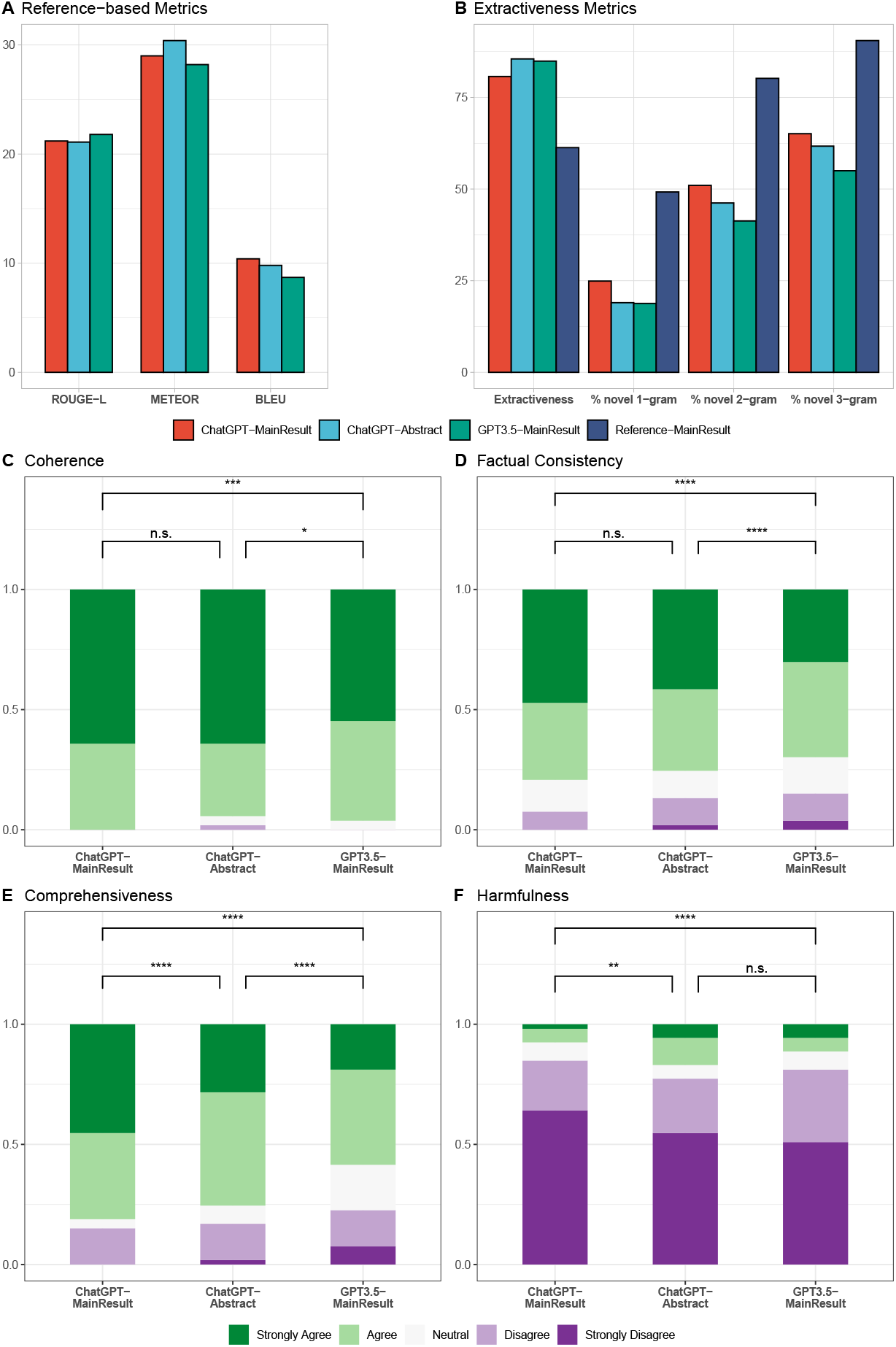
Performance of different summarization systems in automatic and human evaluations. (**A**) Reference-based Metrics (higher scores indicate better summaries). (**B**) Extractiveness Metrics. (**C**) Coherence. (**D**) Factual Consistency. (**E**) Comprehensiveness. (**F**) Harmfulness. Statistical analysis by Mann-Whitney U test (**C-F**), *p-value *≤* 0.05, **p-value *≤* 0.01, ***p-value *≤* 0.001, ****p-value *≤* 0.0001.

### 2.1 Automatic evaluation

To evaluate the quality of the automatically generated summaries, we employed various automatic metrics (Figure 1A), including ROUGE-L [10], METEOR [11], and BLEU [12], comparing them against a reference summary. Their values range from 0.0 to 1.0, with a score of 1.0 indicating the generated summaries are identical to the reference summary. Our findings reveal that all models exhibit similar performance with respect to these automatic metrics. A relatively high ROUGE score demonstrates that these models can effectively capture the key information from the source document. In contrast, a low BLEU score implies that the generated summary is written differently from the reference summary. Consistent METEOR scores across the models suggest that the summaries maintain a similar degree of lexical and semantic similarity to the reference summary.

We also assessed the degree of abstraction by measuring the extractiveness [13] and the percentage of novel n-grams in the summary with respect to the input. Compared to human-written summaries, those generated by LLMs tend to be more extractive, exhibiting significantly lower n-gram novelty (Figure 1B). Notably, ChatGPT-MainResult demonstrates a higher level of abstraction compared to ChatGPT-Abstract and GPT3.5-MainResult, but there remains a substantial gap between ChatGPT-MainResult and human reference. Finally, approximately half of the reviews are from 2022 and 2023, which is after the cutoff date of GPT3.5 (Jun. 2021) and ChatGPT (Sep. 2021). However, we observed no significant difference in quality metrics before and after 2022.

### 2.2 Human evaluation

To obtain a comprehensive understanding of the summarization capabilities of LLMs, we conducted an extensive human evaluation of the model-generated summaries, which goes beyond the capabilities of automatic metrics [14, 15]. Specifically, the lack of standardized terminology of error types for medical evidence summarization necessitated our use of human evaluation to invent new error definitions. Our evaluation methods drew from qualitative methods in grounded theory, which involved open coding of qualitative descriptions of factual inconsistencies, further contributing to the development of error definitions. Additionally, we included a measure of perceived potential for harm, as it is a clinically relevant outcome that automatic metrics are unable to capture. Our evaluation defined summary quality along four dimensions: (1) Coherence; (2) Factual Consistency; (3) Comprehensiveness; and (4) Harmfulness, and the results are presented in Figures 1C-F.

*<u>Coherence</u>* refers to the capability of a summary to create a coherent body of information about a topic through connections between sentences. Figure 1C shows that annotators rated most of the summaries as coherent. Specifically, summaries generated by ChatGPT are more cohesive than those generated by GPT3.5-MainResult (64% vs 55% in Strong agreement).

*<u>Factual Consistency</u>* measures whether the statements in the summary are supported by the source document. As illustrated in Figure 1D, fewer than 10% of summaries produced by ChatGPT-MainResult exhibit factual inconsistency errors, which is significantly lower compared to those generated by other LLM configurations. Medical evidence summaries should be perfectly accurate. To understand the types of factual inconsistency errors that LLMs produce, we categorize these errors into three types of errors using an open coding approach on annotators’ comments (Supplementary Figure 2). Examples can be found in Supplementary Table 1.

*<u>Comprehensiveness</u>* refers to whether a summary contains comprehensive information to support the systematic review. As shown in Figure 1E, both ChatGPT-MainResult and ChatGPT-Abstract provide comprehensive summaries more than 75% of the time, with ChatGPT-MainResult having significantly more summaries annotated as Strongly Agree. In contrast, GPT3.5-MainResult generates noticeably less comprehensive summaries. It would be possible that extending the length of the summary would lead to a more comprehensive summary. However, ChatGPT-MainResult strikes a balance between providing enough information and being concise. The next evaluation was conducted to determine whether the omission of information relevant to the objectives might lead to medical harmfulness.

*<u>Harmfulness</u>* refers to the potential of a summary to cause physical or psychological harm or undesired changes in therapy or compliance due to the misinterpretation of information. Figure 1F shows the error type distributions from summaries that contain harmful information, with ChatGPT-MainResult generating the fewest medically harmful summaries (less than 10%).

Supplement Figure 1 further breaks down the human evaluation for each domain. We observe annotation variations across six different clinical domains, and these variations can be attributed to several factors. (1) The complexity of specific domains or review types may contribute to the observed variability, as some may be less complex than others, making it easier for LLMs to summarize. (2) Domain experts might evaluate the summaries according to their unique internal interpretations of quality metrics. (3) Individual preferences may influence the decision on what key information should be incorporated in the summary.

### 2.3 Human preference

Figure 2 shows the percentage of times humans express a preference for summaries generated by a specific summarization model. Notice that we allow multiple summaries to be selected as the most or least preferred for each source document. As shown in Figure 2A, ChatGPT-MainResult is significantly more preferred among the three LLMs configurations, generating the most preferred summaries approximately half of the time, outperforming its counterparts by a considerable margin. In Figure 2B, we categorize the considerations driving such preference. We find that ChatGPT-MainResult is favored because it produces the most comprehensive summary and includes more salient information. In Figure 2C, the leading reasons for choosing a summary as the least preferred are missing important information, fabricated errors, and misinterpretation errors. This aligns with our finding that ChatGPT-MainResult is the most preferred since it commits the fewest amount of factual inconsistency errors and contains the least harmful or misleading statements.

**Fig. 2:**
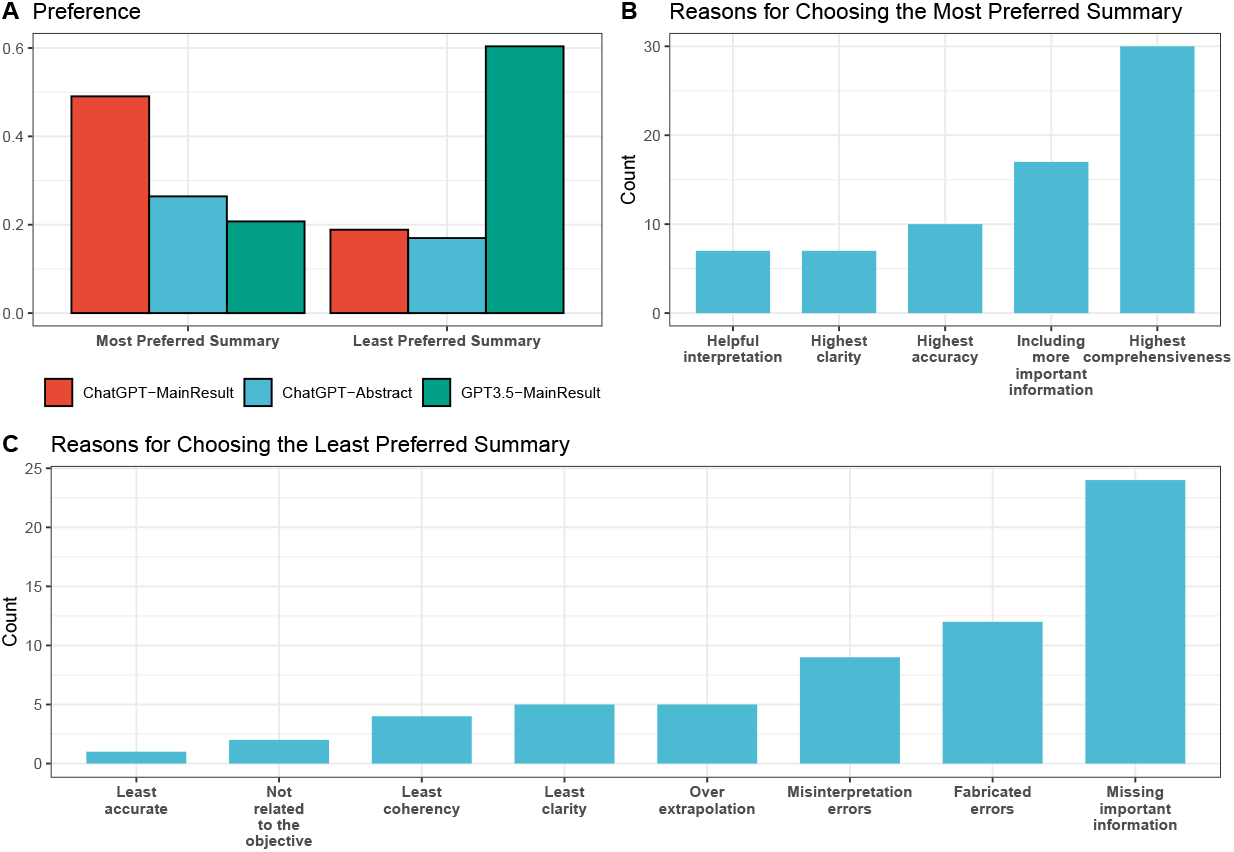
Annotator vote distribution for the most and least preferred summaries (A) and the reasons for choosing them (B and C) across all clinical domains and models.

## 3 Discussions

### 3.1 Are existing automatic metrics well-suited for evaluating medical evidence summaries?

Research has demonstrated that automatic metrics often do not strongly correlate with the quality of summaries [14]. Moreover, there is no off-the-shelf automatic evaluation metric specifically designed to assess the factuality of summaries generated by the most recent summarization systems [7, 16]. We believe this likely extends to the absence of a tailored factuality metric for evaluating medical evidence summaries generated by LLMs as well. In our study, we observed similar results for three model settings when using automatic metrics, which fall short of accurately measuring factual inconsistency, potential for medical harmfulness, or human preference for LLM-generated summaries. Therefore, human evaluation becomes an essential component to properly assess the quality and factuality of medical evidence summaries generated by LLMs at this time, and more effective automatic evaluation methods should be developed for this field.

### 3.2 What causes Factual Inconsistency?

We categorize factual inconsistency errors into three types of errors using an open coding approach on annotators’ comments (Supplementary Figure 2). Examples can be found in Supplementary Table 1.

First, through our qualitative analysis of the annotators’ comments, we discover that auto-generated summaries often contain *<u>Misinterpretation Errors</u>*. These errors can be problematic, as readers might trust the summary’s accuracy without being aware of the potential for falsehoods or distortions. To better understand these errors, we further categorize them into two main sub types. The first is *Contradiction*, which arises when there is a discrepancy between the conclusions drawn from the medical evidence results and the summary. For example, a summary might assert that atypical antipsychotics are effective on psychosis in dementia, whereas the review indicates that the effect is negligible [17]. The other is the *Certainty Illusion*, which occurs when there is an inconsistency in the degree of certainty between the summary and the source document. Such errors may cause summaries to be overly convincing or uncertain, potentially leading readers to rely too heavily on the accuracy of the presented information. For instance, the abstract of a Cochrane Review [18] asserts moderate-certainty evidence that endovascular therapy (ET) plus conventional medical treatment (CMT) compared to CMT alone causes a higher risk of short-term stroke and death. However, we found that the generated summary conveys low-quality confidence.

*<u>Fabricated Errors</u>* arise when a statement appears in a summary, but no evidence from the source document can be found to support or refute the statement. For instance, a summary states that exercise could enhance satisfaction and quality of life for patients with chronic neck pain, but the review does not mention those two outcomes for patients [19]. Interestingly, in our human evaluation, we did not find ChatGPT producing any fabricated errors.

Finally, *<u>Attribute Errors</u>* refer to any errors on non-key elements in the review question (i.e., Patient/ Problem, Intervention, Comparison, and Outcome) and may arise in summaries under three circumstances: (a) *Fabricated Attribute*: This error occurs when a summary incorporates an attribute for a specific symptom or outcome that is not referenced in the source document. For example, a review [18] draws conclusions about a population with intracranial artery stenosis (ICAS), but the summary states the population with recent symptomatic severe ICAS, where “recent” and “severe” cannot be inferred from the document and would impact one’s interpretation of the review; (b) *Omitted Attribute*: This error occurs when a summary neglects an attribute for a specific symptom or outcome, such as neglecting to specify the subtype of dementia discussed in the review, leading to overgeneralization of the conclusion [17]; and (c) *Distorted Attribute*: This error occurs when the specified attribute is incorrect, like stating four trials are included in the study while the source document indicates that only two trials are included [20].

We observed that ChatGPT-MainResult has the lowest proportion of all three types of errors when compared to the other two model configurations. Moreover, it is important to note that LLMs generally generate summaries with few fabricated errors. This is a promising finding, as it is crucial for generated statements to be supported by source documents. However, LLMs do display a noticeable occurrence of attribute errors and misinterpretation errors, with ChatGPT-Abstract and GPT3.5-MainResult displaying a higher incidence of the latter. Drawing inaccurate conclusions or conveying incorrect certainty regarding evidence could lead to medical harm as shown in later sections.

### 3.3 What causes Medical Harmfulness?

We further identify three reasons that could potentially cause medical harmfulness: misinterpretation errors, fabricated errors, and incomprehensiveness (such as missing Population, Intervention, Comparison, Outcome (PICO) elements). Notably, we did not find any instances of medical harmfulness resulting from attribute errors in the summaries we analyzed. However, given the limited number of summaries we examined, we cannot definitively conclude that attribute errors could never cause harm. Our study suggests that medical harmfulness caused by LLMs is mainly due to misinterpretation errors and incomprehensiveness. Although our human evaluation showed that LLMs tend to make relatively few fabricated errors when completing our tasks, we cannot exclude the possibility that such errors could lead to harmful consequences. However, not all summaries with these errors would bring medical harm. For example, although the summary makes a significant error by misspecifying the number of trials in a study [20], our domain experts do not think this could bring medical harm.

### 3.4 How do human-generated summaries compare to LLM-generated summaries?

We observe that human-generated summaries contain a higher proportion (28%) of fabricated errors, resulting in more factual inconsistency and potential for harmfulness in human references. However, it is essential to approach this finding with caution, as our human evaluation on human-generated summaries relies solely on the abstracts of Cochrane Reviews as a proxy. There is a possibility that statements deemed to contain fabricated errors could, in fact, be validated by other sections within the fulllength Cochrane Review. For example, the severity of ICAS of the studied population (see the example for Fabricated Attribute) is not mentioned in the abstract of the review but it is mentioned in the Results section of the whole review. Therefore, we decide to exclude the human reference from our comparison figures. Nevertheless, despite these errors, human-generated summaries are still preferred (34%) compared to ChatGPT-Abstract (26%) and GPT3.5-MainResult (21%). It is worth noting that human-generated summaries may contain valuable interpretations of reviews, which account for why they are chosen as the best summaries. However, it is important to avoid over-extrapolating from the source document, as this could lead to less desirable outcomes (as illustrated in Figures 2B and 2C).

### 3.5 Does providing longer input lead to better summaries generated by LLMs?

It is important to emphasize that the main difference between ChatGPT-MainResult and ChatGPT-Abstract is that the latter generates summaries based on the entire abstract. Our findings show that having longer text actually negatively impacts Chat-GPT’s capability to identify and extract the most pertinent information, as evidenced by the lower comprehensiveness. Furthermore, a longer context leads to an increased likelihood of ChatGPT making factual inconsistency errors and generating summaries that are more misleading. These factors combined make ChatGPT-Abstract less preferred compared to ChatGPT-MainResult in human evaluation.

### 3.6 How can we automatically detect factual inconsistency and improve summaries?

Given that the types of errors made by the most recent summarization systems are constantly evolving [16], future factuality and harmfulness evaluation should be adaptable to these shifting targets. One possible approach is to leverage the power of LLMs to identify potential errors within summaries. However, effectively identifying the most important information from long contexts and making high-quality summaries remains a challenging task for LLMs that we have evaluated. Methods such as segment-then-summarize [21] and extract-then-abstract [21] for handling summarizing long context are shown to not work well for zero-shot LLM [7].

Furthermore, the presence of non-textual data, such as tables and figures in Cochrane Reviews, may increase the complexity of the summarization task. To address these challenges and improve the quality of summaries, future work could explore and evaluate the efficacy of GPT-4 in summarizing reviews with longer contexts and multiple modalities, while also incorporating techniques for detecting factuality inconsistencies and medical harmfulness.

### 3.7 Limitations

Our evaluation of ChatGPT and GPT-3.5 is based on a semi-synthetic task, which involves summarizing Cochrane Reviews using only their abstracts or part of the abstracts. A more genuine task here would be a multi-document summarization setting that involves summarizing all relevant study reports within a review addressing specific research questions. The rationale behind this choice is three-fold. First, the abstracts of Cochrane Reviews is a stand-alone documents [9] that should not contain any information that is not in the main body of the review, and the overall messages should be consistent with the conclusions of the review. Second, abstracts of Cochrane Reviews are freely available and may be the only source readers can assess the review results. Finally, we need to accommodate the input length constraints of large language models, as the full Cochrane Review would surpass their capacity. As a result, our experiment provides an assessment of these LLMs’ ability to summarize medical evidence under a modified summarization framework. Our rigorous systematic evaluation finds that ChatGPT tends to generate less factually accurate summaries when conditioned on the entire abstract, which could potentially indicate that the model may be susceptible to distraction from irrelevant information within longer contexts. This finding raises concerns about the model’s effectiveness when presented with the full scope of a Cochrane Review, and suggests that it may not perform optimally in such scenarios.

Secondly, the prompt in this study is adapted from previous work [7]. Given the lack of a systematic method for searching over the prompt space, it is conceivable that future studies could potentially discover more effective prompts.

Our evaluation of LLM-generated summaries focused on six clinical domains, with one designated expert assigned to each domain. Such evaluation requires domain knowledge, making it difficult for non-experts to carry out the evaluation. This constraint limits the total amount of summaries we are able to annotate. Further, we chose to use only the abstracts of Cochrane Reviews to evaluate human-generated summaries (*Author’s Conclusions* section) since examining the entire Cochrane Review is a time-consuming process. Therefore, it is possible that some of the errors identified in human reference summaries may actually be substantiated by other sections of the full-length review.

## 4 Methods

### 4.1 Materials

A Cochrane Review is a systematic review of scientific evidence that aims to provide a comprehensive summary of all relevant studies related to a specific research question. Reviews follow a rigorous methodology, which includes a comprehensive search for relevant studies, the critical appraisal of study quality, and the synthesis of study findings. The primary objective of Cochrane Reviews is to provide an unbiased and comprehensive summary and meta analysis of the available evidence, to help healthcare professionals make informed decisions about the most effective treatment options.

In this work, we utilized Cochrane Reviews extracted from the Cochrane Library, which is a large database that provides high-quality and up-to-date information about the effects of healthcare interventions. It covers a diverse range of healthcare topics, and our study focuses on six specific topics drawn from this resource – Alzheimer’s disease, Kidney disease, Esophageal cancer, Neurological conditions, Skin disorders, and Heart failure. In particular, we have collected approximately ten most-recent reviews on each topic. Each review was verified by domain experts to ensure that they have important research objectives. Table 1 shows the basic summary statistics.

We focus on the abstracts of Cochrane Reviews in our study, which can be read as stand-alone documents. Each abstract includes the Background, Objectives of the review, Search methods, Selection criteria, Data collection and analysis, Main results, and Author’s conclusions.

### 4.2 Experimental setup

In this study, we aim to evaluate the zero-shot performance of summarizing systematic reviews using two OpenAI-developed models: GPT-3.5 (text-davinci-003) and Chat-GPT. GPT-3.5, or InstructGPT, is built upon the GPT-3 model but has undergone further training using reinforcement learning with a human feedback procedure with the goal of providing better outputs preferred by humans. ChatGPT has gathered significant attention due to its ability to generate high-quality and human-like responses to conversational text prompts. Despite its impressive capabilities, it remains unclear whether ChatGPT can generalize and perform high quality zero-shot summarization of medical evidence reviews. Therefore, we seek to investigate the comparative performance of ChatGPT and GPT-3.5 in summarizing systematic reviews of medical evidence data.

To evaluate the capabilities of the models, we have designed two distinct setups for input. In the first setup, the models take the whole abstract except for the *Author’s Conclusions* as the input (ChatGPT-Abstract). The second setup relies on taking both the *objectives* and the *Main results* sections of the abstract as the model input (ChatGPT-MainResult and GPT3.5-MainResult). The objective section outlines the specific research question in the PICO (Population, Intervention, Comparison, Outcome) formulation that the review aims to address, while the Main results section provides a summary of the results of the studies included in the review, including key outcomes and any statistical data, while highlighting the strengths and limitations of the evidence.

In both settings, we use *[input*] + “*Based on the Objectives, summarize the above systematic review in four sentences*” to prompt the model to perform summarization, where we emphasize the purpose of the summary by providing the *Objectives* section. We use the *Author’s Conclusions* section as the human reference (see explanations in the Introduction) and compare it against the models’ generated outputs.

### 4.3 Automatic Evaluation Metrics

We select several metrics that have been widely used in text generation and summarization. ROUGE-L [10] measures the overlap between the generated summary and the reference summary, focusing on the recall of the n-grams. METEOR [11] measures the harmonic mean of unigram precision and recall, based on stemming and synonym matching. BLEU [12] measures the overlap between the generated summary and the reference summary, focusing on the precision of the n-grams. In addition, we selected two reference-free metrics. Extractiveness [13] measures the percentage of words in a summary that is from the source document. The percentage of novel n-grams signifies the proportion of n-grams in the summary that differ from the original source document.

### 4.4 Design of the Human Evaluation

We systematically evaluate the quality of generated summaries via human evaluation. We propose to evaluate summary quality along several dimensions: (1) Factual consistency; (2) Medical harmfulness; (3) Comprehensiveness; and (4) Coherence. These dimensions have been previously identified and serve as essential factors in evaluating the overall quality of generated summaries [15, 22, 23]. Factual consistency measures whether the statements in the summary are supported by the systematic review. Medical harmfulness refers to the potential of a summary that leads to physical or psychological harm or unwanted changes in therapy or compliance due to the misinterpretation of information. Comprehensiveness evaluates whether a summary contains sufficient information to cover the objectives of the systematic review. Coherence refers to the ability of a summary to build a coherent body of information about a topic through sentence-to-sentence connections.

To assess the quality of the generated summaries, we include six domain experts, with each annotating summaries for a specific topic. During the annotation process, participants are presented with the whole abstract of the systematic review, along with four summaries: (1) the *Authors’ conclusion* section; (2) ChatGPT-MainResult; (3) ChatGPT-Abstract; and (4) GPT3.5-MainResult. The order in which the summaries are presented is randomized to minimize potential order effects during the evaluation process. We utilize a 5-point Likert scale for the evaluation of each dimension. If the summary received a low score on any of the dimensions, we further asked participants to explain the reason for the low score in a provided text box for each dimension. This approach enables us to perform a qualitative analysis of the responses and identify common themes to define a terminology of error types for medical evidence summarization where none exists. In addition to evaluating the quality of the summaries, we also asked participants to indicate their most and least preferred summaries and to provide reasons for their choices. This approach enables us to identify specific subcategories of reasons and gain insights into the potential of using model-generated summaries to assist in completing the systematic review process.

The 5-point Likert scales between models were assessed by the Mann-Whitney U test [24]. The response categories of a 5-point Likert item are coded 1 to 5 which were used as numerical scores in the Mann-Whitney U test for differences. The p-value reflects if the responses of the summaries generated by two models are different, assuming the null hypothesis means there is no difference between the results generated by the two models. We used 1,000 bootstrap samples to obtain a distribution of the Likert scales and reported p-values.

## Data Availability

All data produced in the present study are available upon reasonable request to the authors

https://www.cochranelibrary.com

## Acknowledgments

This work was supported by the National Library of Medicine (NLM) of the National Institutes of Health (NIH) under grant number 4R00LM013001 and 5R01LM009886, NIH Bridge2AI (OTA-21-008), National Science Foundation under grant numbers 2145640, 2019844, and 2303038, and Amazon Research Award. The content is solely the responsibility of the authors and does not necessarily represent the official views of the NIH and NSF.

## Supplementary materials

**Table 1:** An Overview of Error Types.

| Error type | Definition | Example |  | Consequence |
| --- | --- | --- | --- | --- |
|  |  | Source document | Summary |  |
| Misinterpretation errors |  |  |  |  |
| Contradiction | Discrepancy between the conclusions drawn from the medical evidence results and the summary | The effect of atypical antipsychotic on psychosis in dementia is negligible | Atypical antipsychotics are effective on psychosis in dementia | Potential for falsehoods |
| Certainty Illusion | Inconsistency in the degree of certainty between the summary and the source document | There is low-quality confidence that endovascular therapy (ET) plus conventional medical treatment (CMT) compared to CMT alone causes a higher risk of short-term stroke and death | There is moderate-certainty evidence that ET plus CMT compared to CMT alone causes a higher risk of short-term stroke and death | Overly convincing or uncertain summary |
| Fabricated errors |  |  |  |  |
|  | No evidence from the source document can be found to support or refute the statement | Two outcomes "help enhance satisfaction and quality of life" are not mentioned in the review | Exercise could help reduce pain and disability and help enhance satisfaction and quality of life for patients with chronic neck pain | Lack of evidence |
| Attribute errors |  |  |  |  |
| Fabricated Attribute | Incorporate an attribute for a specific symptom or outcome that is not referenced in the source document | Population with intracranial artery stenosis (ICAS) | Population with recent symptomatic severe ICAS, where "recent" and "severe" cannot be inferred from the source | Misinterpretation of the review |
| Omitted Attribute | Neglect an attribute for a specific symptom or outcome | Specify the subtype of dementia | Not specify the subtype of dementia | Overgeneralization of the conclusion |
| Distorted Attribute | Generate incorrect attribute | Two trials are included in the study | Four trials are included in the study | Misinformation of the study |

**Fig. 1:**
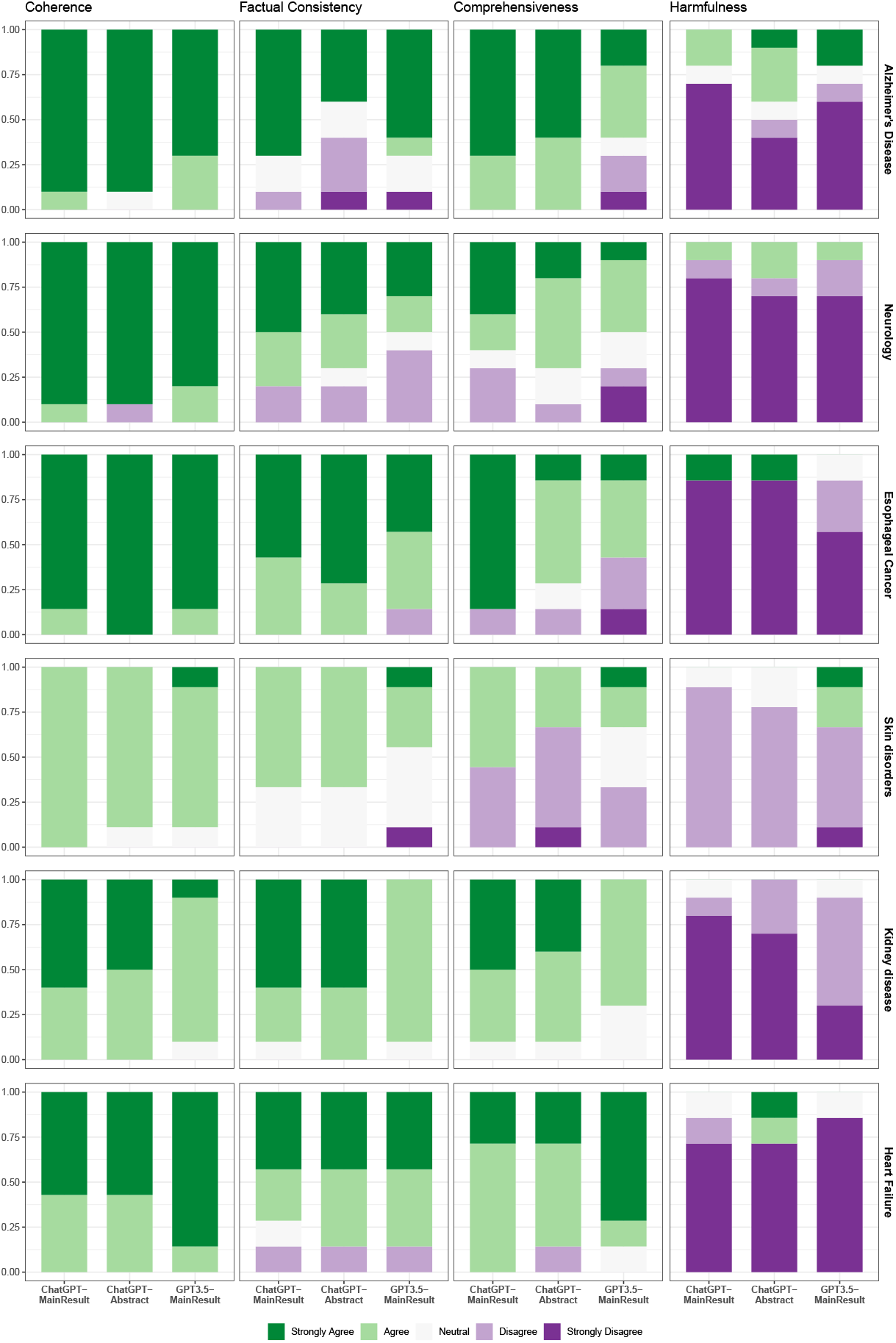
Annotator vote distribution for (A) Coherence, (B) Factual Consistency, (C) Comprehensiveness, and (D) Harmfulness of summaries generated by different summarization systems in 6 clinical domains.

**Fig. 2:**
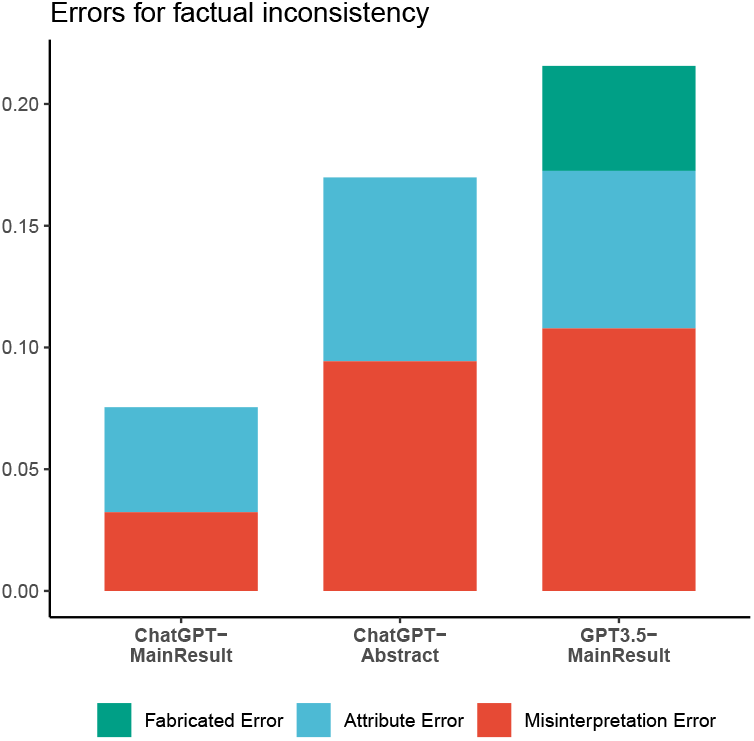
Statistics of errors for factual inconsistency across models.

## References

[1] Chowdhery, A., Narang, S., Devlin, J., Bosma, M., Mishra, G., Roberts, A., Barham, P., Chung, H.W., Sutton, C., Gehrmann, S., Schuh, P., Shi, K., Tsvyashchenko, S., Maynez, J., Rao, A., Barnes, P., Tay, Y., Shazeer, N., Prabhakaran, V., Reif, E., Du, N., Hutchinson, B., Pope, R., Bradbury, J., Austin, J., Isard, M., Gur-Ari, G., Yin, P., Duke, T., Levskaya, A., Ghemawat, S., Dev, S., Michalewski, H., Garcia, X., Misra, V., Robinson, K., Fedus, L., Zhou, D., Ippolito, D., Luan, D., Lim, H., Zoph, B., Spiridonov, A., Sepassi, R., Dohan, D., Agrawal, S., Omernick, M., Dai, A.M., Pillai, T.S., Pellat, M., Lewkowycz, A., Moreira, E., Child, R., Polozov, O., Lee, K., Zhou, Z., Wang, X., Saeta, B., Diaz, M., Firat, O., Catasta, M., Wei, J., Meier-Hellstern, K., Eck, D., Dean, J., Petrov, S., Fiedel, N.: PaLM: Scaling language modeling with pathways. arXiv preprint arXiv:2204.02311 (2022) arXiv:2204.02311 [cs.CL]

[2] Wei, J., Wang, X., Schuurmans, D., Bosma, M., Ichter, B., Xia, F., Chi, E., Le, Q., Zhou, D.: Chain-of-thought prompting elicits reasoning in large language models. In: Koyejo, S., Mohamed, S., Agarwal, A., Belgrave, D., Cho, K., Oh, A. (eds.) Advances in Neural Information Processing Systems, vol. 35, pp. 24824–24837. Curran Associates, Inc., ??? (2022)

[3] Kojima, T., Gu, S.s., Reid, M., Matsuo, Y., Iwasawa, Y.: Large language models are Zero-Shot reasoners. In: Koyejo, S., Mohamed, S., Agarwal, A., Belgrave, D., Cho, K., Oh, A. (eds.) Advances in Neural Information Processing Systems, vol. 35, pp. 22199–22213. pnCurran Associates, Inc., ??? (2022)

[4] Brown, T., Mann, B., Ryder, N., Subbiah, M., Kaplan, J.D., Dhariwal, P., Neelakantan, A., Shyam, P., Sastry, G., Askell, A., Agarwal, S., Herbert-Voss, A., Krueger, G., Henighan, T., Child, R., Ramesh, A., Ziegler, D., Wu, J., Winter, C., Hesse, C., Chen, M., Sigler, E., Litwin, M., Gray, S., Chess, B., Clark, J., Berner, C., McCandlish, S., Radford, A., Sutskever, I., Amodei, D.: Language models are Few-Shot learners. In: Larochelle, H., Ranzato, M., Hadsell, R., Balcan, M.F., Lin, H. (eds.) Advances in Neural Information Processing Systems, vol. 33, pp. 1877–1901. Curran Associates, Inc., ??? (2020)

[5] Ouyang, L., Wu, J., Jiang, X., Almeida, D., Wainwright, C., Mishkin, P., Zhang, C., Agarwal, S., Slama, K., Ray, A., Schulman, J., Hilton, J., Kelton, F., Miller, L., Simens, M., Askell, A., Welinder, P., Christiano, P.F., Leike, J., Lowe, R.: Training language models to follow instructions with human feedback. In: Koyejo, S., Mohamed, S., Agarwal, A., Belgrave, D., Cho, K., Oh, A. (eds.) Advances in Neural Information Processing Systems, vol. 35, pp. 27730–27744. Curran Associates, Inc., ??? (2022)

[6] OpenAI: Introducing ChatGPT. ChatGPT. Accessed: 2023-4-15 (2023)

[7] Goyal, T., Li, J.J., Durrett, G.: News summarization and evaluation in the era of GPT-3. arXiv preprint arXiv:2209.12356 (2022) arXiv:2209.12356 [cs.CL]

[8] Gao, C.A., Howard, F.M., Markov, N.S., Dyer, E.C., Ramesh, S., Luo, Y., Pearson, A.T.: Comparing scientific abstracts generated by ChatGPT to original abstracts using an artificial intelligence output detector, plagiarism detector, and blinded human reviewers. bioRxiv (2022)

[9] Beller, E.M., Glasziou, P.P., Altman, D.G., Hopewell, S., Bastian, H., Chalmers, I., Gøtzsche, P.C., Lasserson, T., Tovey, D., PRISMA for Abstracts Group: PRISMA for abstracts: reporting systematic reviews in journal and conference abstracts. PLoS Med. 10(4), 1001419 (2013) https://doi.org/10.1371/journal.pmed.1001419

[10] Lin, C.-Y.: ROUGE: a package for automatic evaluation of summaries. In: Text Summarization Branches Out: Proceedings of the ACL-04 Workshop, vol. 8, pp. 1–8. Barcelona, Spain, ??? (2004)

[11] Banerjee, S., Lavie, A.: METEOR: an automatic metric for MT evaluation with improved correlation with human judgments. In: Proceedings of the ACL Workshop on Intrinsic and Extrinsic Evaluation Measures for Machine Translation And/or Summarization, pp. 65–72 (2005)

[12] Papineni, K., Roukos, S., Ward, T., Zhu, W.-J.: BLEU: a method for automatic evaluation of machine translation. In: Proceedings of the 40th Annual Meeting on Association for Computational Linguistics. ACL ‘02, pp. 311–318. Association for Computational Linguistics, USA (2002)

[13] Grusky, M., Naaman, M., Artzi, Y.: Newsroom: A dataset of 1.3 million summaries with diverse extractive strategies. In: Proceedings of the 2018 Conference of the North American Chapter of the Association for Computational Linguistics: Human Language Technologies, Volume 1 (Long Papers), pp. 708–719. Association for Computational Linguistics, Stroudsburg, PA, USA (2018). https://doi.org/10.18653/v1/n18-1065

[14] Fabbri, A.R., Kryścińki, W., McCann, B., Xiong, C., Socher, R., Radev, D.: SummEval: Re-evaluating summarization evaluation. Trans. Assoc. Comput. Linguist. 9, 391–409 (2021)

[15] Tang, L., Kooragayalu, S., Wang, Y., Ding, Y., Durrett, G., Rousseau, J.F., Peng, Y.: EchoGen: Generating conclusions from echocardiogram notes. In: Proceedings of the 21st Workshop on Biomedical Language Processing, pp. 359–368. Association for Computational Linguistics, Dublin, Ireland (2022)

[16] Tang, L., Goyal, T., Fabbri, A.R., Laban, P., Xu, J., Yahvuz, S., Kryścińki, W., Rousseau, J.F., Durrett, G.: Understanding factual errors in summarization: Errors, summarizers, datasets, error detectors. arXiv preprint arXiv:2205.12854 (2022) arXiv:2205.12854 [cs.CL]

[17] Mühlbauer, V., Möhler, R., Dichter, M.N., Zuidema, S.U., Köpke, S., Luijendijk, H.J.: Antipsychotics for agitation and psychosis in people with alzheimer’s disease and vascular dementia. Cochrane Database Syst. Rev. 12(12), 013304 (2021) https://doi.org/10.1002/14651858.CD013304.pub2

[18] Luo, J., Wang, T., Yang, K., Wang, X., Xu, R., Gong, H., Zhang, X., Wang, J., Yang, R., Gao, P., Ma, Y., Jiao, L.: Endovascular therapy versus medical treatment for symptomatic intracranial artery stenosis. Cochrane Database Syst. Rev. 2, 013267 (2023) https://doi.org/10.1002/14651858.CD013267.pub3

[19] Gross, A., Kay, T.M., Paquin, J.-P., Blanchette, S., Lalonde, P., Christie, T., Dupont, G., Graham, N., Burnie, S.J., Gelley, G., Goldsmith, C.H., Forget, M., Hoving, J.L., Brønfort, G., Santaguida, P.L., Cervical Overview Group: Exercises for mechanical neck disorders. Cochrane Database Syst. Rev. 1(1), 004250 (2015)

[20] Kamo, T., Wada, Y., Okamura, M., Sakai, K., Momosaki, R., Taito, S.: Repetitive peripheral magnetic stimulation for impairment and disability in people after stroke. Cochrane Database Syst. Rev. 9(9), 011968 (2022) https://doi.org/10.1002/14651858.CD011968.pub4

[21] Zhang, Y., Ni, A., Mao, Z., Wu, C.H., Zhu, C., Deb, B., Awadallah, A., Radev, D., Zhang, R.: SummN : A Multi-Stage summarization framework for long input dialogues and documents. In: Proceedings of the 60th Annual Meeting of the Association for Computational Linguistics (Volume 1: Long Papers), pp. 1592– 1604. Association for Computational Linguistics, Dublin, Ireland (2022). https://doi.org/10.18653/v1/2022.acl-long.112

[22] Singhal, K., Azizi, S., Tu, T., Sara Mahdavi, S., Wei, J., Chung, H.W., Scales, N., Tanwani, A., Cole-Lewis, H., Pfohl, S., Payne, P., Seneviratne, M., Gamble, P., Kelly, C., Scharli, N., Chowdhery, A., Mansfield, P., Arcas, B., Webster, D., Corrado, G.S., Matias, Y., Chou, K., Gottweis, J., Tomasev, N., Liu, Y., Rajkomar, A., Barral, J., Semturs, C., Karthikesalingam, A., Natarajan, V.: Large language models encode clinical knowledge. arXiv preprint arXiv:2212.13138 (2022) arXiv:2212.13138 [cs.CL]

[23] Jeblick, K., Schachtner, B., Dexl, J., Mittermeier, A., Stüber, A.T., Topalis, J., Weber, T., Wesp, P., Sabel, B., Ricke, J., Ingrisch, M.: ChatGPT makes medicine easy to swallow: An exploratory case study on simplified radiology reports. arXiv preprint arXiv:2212.14882 (2022) arXiv:2212.14882 [cs.CL]

[24] Clason, D., Dormody, T.: Analyzing data measured by individual likert-type items. J. Agric. Educ. 35(4), d31–35 (1994)

